# A Systematic Review and Meta-analysis of Therapeutic options against SARS-CoV-2

**DOI:** 10.1101/2020.05.20.20108365

**Authors:** Viveksandeep Thoguluva Chandrasekar, Bhanuprasad Venkatesalu, Harsh K. Patel, Marco Spadaccini, Jacob Manteuffel, Mayur Ramesh

**Author notes:** Corresponding author: Viveksandeep Thoguluva Chandrasekar University of Kansas Medical Center, Kansas City, Kansas 66160, United States, Email id.

## Abstract

**Importance:** Treatment options for Severe acute respiratory syndrome-related coronavirus-2 (SARS-CoV-2) are limited with no clarity on the efficacy and safety profiles.

**Objective:** To assess if the effect estimate of any intervention improves the outcomes and safety profile.

**Data sources:** PubMed, Embase, Cochrane Central were searched from December 1, 2019 to May 11, 2020.

**Study selection:** Any prospective/retrospective clinical study on SARS-CoV-2 patients ≥18 years of age with report on therapeutic interventions.

**Data synthesis and extraction:** Data was screened and extracted by two independent investigators.

**Main outcomes and measures:** The primary outcome was all-cause in-hospital mortality. The secondary outcomes were rates of mechanical ventilation, viral clearance, adverse events, discharge, progression to severe disease, median time for clinical recovery and anti-viral clearance. Pooled rates and odds ratios (OR) were calculated.

**Results:** A total of 29 studies with 5207 participants were included in the analysis. The pooled all-cause in-hospital mortality rate was 12.8% (95%CI: 8.1%-17.4%) in intervention arm. There was no significant difference in mortality between both arms overall (OR: 1.36, 95% CI: 0.97-1.89). The mortality was significantly higher in the Hydroxychloroquine (HCQ) group compared to control: (1.86, 95% CI: 1.38 – 2.50). The need for mechanical ventilation in patients with mild-moderate disease was 13.5% vs 9.8% in intervention and control groups, with no significant difference (OR: 1.58, 95% CI: 0.60 – 4.15).The median duration for viral clearance in the intervention arm was 6.1 (IQR: 4.3 – 8.8) days and control arm was 9 (IQR: 4.5 – 14) days, with no significant difference between the groups (p = 0.37). There was no significant difference between pooled adverse event rates in intervention and control groups: 34% vs 29.5% (OR: 1.44, 95% CI: 0.70 – 2.94), respectively. However, incidence of adverse events was significantly higher in HCQ sub-group (OR: 3.88, 95% CI: 1.60 – 9.45, I^2^ = 0%). There was no significant difference in other secondary outcomes.

**Conclusion and relevance:** The use of hydroxychloroquine was associated with increased mortality and adverse event rates. No other therapeutic intervention including Lopinavir/Ritonavir, Remdesivir or Tocilizumab seem to alter the natural course of the disease. There is a further need for well-designed randomized clinical trials.

**Article summary line:** The use of hydroxychloroquine was associated with increased mortality and adverse event rates in Severe acute respiratory syndrome-related coronavirus-2 infection and other therapeutic interventions did not show any difference in outcomes

## Introduction

Severe acute respiratory syndrome-related coronavirus-2(SARS-CoV-2) is the 7^th^ virus of the coronavirus family known to infect humans(1). By March, the World health Organization had declared SARS-CoV-2 as a pandemic, the third pandemic in the 21^st^ century after the SARS outbreak in 2003 and H1N1 influenza in 2009. SARS-CoV-2 tends to cause a plethora of symptoms with fever, cough, myalgia, fatigue, loss of taste, appetite and diarrhea to name a few. It is also known to affect multiple organ systems leading to acute respiratory distress syndrome, encephalitis, myocarditis, hepatitis, acute kidney injury and hypercoagulable state leading to stroke and pulmonary embolism. The COVID-19 disease caused by SARS-CoV-2 can be classified as mild, moderate, severe and critical disease based on clinical, imaging and laboratory parameters(2).The natural history of the disease is such that most patients typically have mild disease with spontaneous resolution of symptoms by 10-14 days needing symptomatic management and home self-quarantine. Elderly population, as well as patients with medical co-morbidities are at higher risk of developing moderate to severe disease(3). As per the Chinese Center for Disease Control and Prevention data in a cohort of 72,314 patients, clinical deterioration tends to typically occur in the second week of onset of symptoms with need for hospitalization and close monitoring in 14% of patients and around 5% of patients require invasive ventilation (4). Several therapeutic interventions like Hydroxychloroquine (HCQ), Chloroquine, Remdesivir, Corticosteroids, Tocilizumab and convalescent plasma therapy have been attempted, but currently there is no known intervention that has reduced mortality in COVID-19 patients. These questions bring into focus the need of a comprehensive systematic review of the published literature to collate the available evidence. The aim of this systematic review and meta-analysis is to assess if any intervention provides mortality benefit, other clinically relevant outcomes and also ascertain the safety profile.

## Methods

This systematic review was performed as per the Preferred Reporting Items for Systematic Reviews and Meta-Analyses (PRISMA) recommendations(5).The protocol is provided as **Appendix 1**. Institutional review board approval was not required for this study since no patient identifiers were disclosed.

### Data sources

A systematic electronic search was performed in PubMed/MEDLINE, Embase, Cochrane Central, Google Scholar, MedRxiv databases to identify published and pre-published studies reporting outcomes related to interventions for SARS-CoV-2 infection, from December 1, 2019 to May 11, 2020. The Medical Subject Heading/ Entree terms is provided in **Appendix 2**. An independent review of the abstracts and full paper articles was done (VT and BV). The duplicates were removed and the titles of articles were evaluated. The full-length papers of the shortlisted articles were assessed for the eligibility criteria. The articles that fulfilled the inclusion criteria were shortlisted for final systematic review. The included study references were cross-searched for additional studies. The articles were reviewed independently by two authors (VT and BV) and any disagreement was resolved by consensus with a third author (MR). Reasons for excluding studies were documented.

### Eligibility criteria

The inclusion criteria were as follows: 1) Studies reporting outcomes for treatment in SARS-CoV-2 infection 2) All studies including randomized controlled trials (RCTs), prospective, retrospective and case series 3) Full length studies 4) Patients more than 18 years of age. Exclusion criteria were: 1) Pre-clinical studies, epidemiological and descriptive studies without intervention for SARS-CoV-2 patients 2) Abstracts.

### Data extraction and quality assessment

The data was extracted by two authors independently into pre-defined forms. The following data was extracted from the studies: first author, mean age, study design, number of patients, gender, rates of: mortality, clinical improvement, mechanical ventilation, progression to severe disease, viral clearance, discharge and adverse events. Data for both intervention and control arms (for available studies) were extracted separately. Quality assessment was performed only for RCTs as most of the other studies were retrospective in nature with short hospital courses for duration of treatment. Cochrane risk bias tool was used for study quality assessment forRCTs(6).

### Definitions and Outcomes

The definitions of outcomes that were assessed is provided as **Appendix 3**. The intervention arm consisted of patients receiving the drug or the therapeutic intervention while the control arm patients received standard of care treatment for SARS-CoV-2 without a specific intervention. The primary outcomes were the all-cause mortality in the intervention arm and in comparison, with control arm. The secondary outcomes were rates of clinical recovery, need for mechanical ventilation, viral clearance, radiological improvement, discharge and adverse events in intervention arm and comparison with control arm. Median duration for viral clearance and clinical recovery were also calculated from available studies. Number needed to treat (NNT) and number needed to harm (NNH) were defined as the number of patients who needed to be treated to provide benefit or harm in at least 1 patient, comparing intervention and control arms for respective outcomes.

### Statistical analysis

Percentages for categorical variables and median with interquartile range (IQR) for continuous variables were presented. Differences in medians were calculated using the Mann-Whitney-Wilcoxon test. Proportions with pooled rates with 95% confidence intervals (CI) were calculated for individual arms. Odds ratios (OR) comparing with control arm was reported with 95% CI and p-value <0.05 was considered statistically significant. Random effects model described by DerSimonian and Laird were used for analysis. Corresponding forest plots were constructed for both primary and secondary outcomes. NNT and NNH were calculated using the inverse of the differences in benefit or harm between the intervention and control arms for the respective outcomes. Study heterogeneity was assessed using Inconsistency index (I^2^-statistic) with low, moderate, substantial and considerable heterogeneity indicated by I^2^-value of 0-30%, 31%-60%, 61%-75% and 76%-100%, respectively. All analyses were performed using statistical softwares Open Meta analyst (CEBM, Brown University, Rhode Island, USA) and Review Manager Version 5.3 (The Nordic Cochrane Center, Copenhagen, Denmark). Sub-group analyses were performed for the following, when data was available and also to address heterogeneity in primary outcome if present: 1) Intervention specific, 2) Disease severity specific, 3) RCTs only.

## Results

### Study search and study characteristics

The literature search resulted in 3664 articles, of which 65 articles underwent full review and 29 were included in the final analysis (**Figure 1**)(3, 7-34). Among included studies, 19 were performed in China, 4 in France, 4 in USA, 1 in Brazil and 1 in South Korea. Eight studies were RCTs, 4 were prospective studies and the remaining 17 were retrospective studies. Fifteen studies were published and the remaining were pre-published. Seventeen studies had a drug or intervention being tested with a control group for comparison. For intervention, 12 studies used HCQ based treatment (2 studies had azithromycin along with HCQ in same arm, 3 had azithromycin in separate arm and 1 study was comparison of HCQ with Lopinavir/ Ritonavir), 5 studies used antiviral agents (2 studies with Lopinavir/ Ritonavir, 1 with Baloxavir/ Marboxil and Favipravir and 2 with Remdesivir), 2 were Tocilizumab based single arm studies, 5 used corticosteroids (3 with control arm) and 5 studies were single-arm plasma therapy based. There were 3,624 patients in the intervention arm (mean age: 55.9 ± 8.4 years, 62% males) and 1,583 patients (mean age: 52.5 ± 8.5 years, 60.7% males) in the control arm. The median duration of follow-up was 14 days (IQR: 9 – 24.5) and the range was 6-32 days across all studies. The demographics and study characteristics have been provided in **Table 1**.

**Figure 1.**
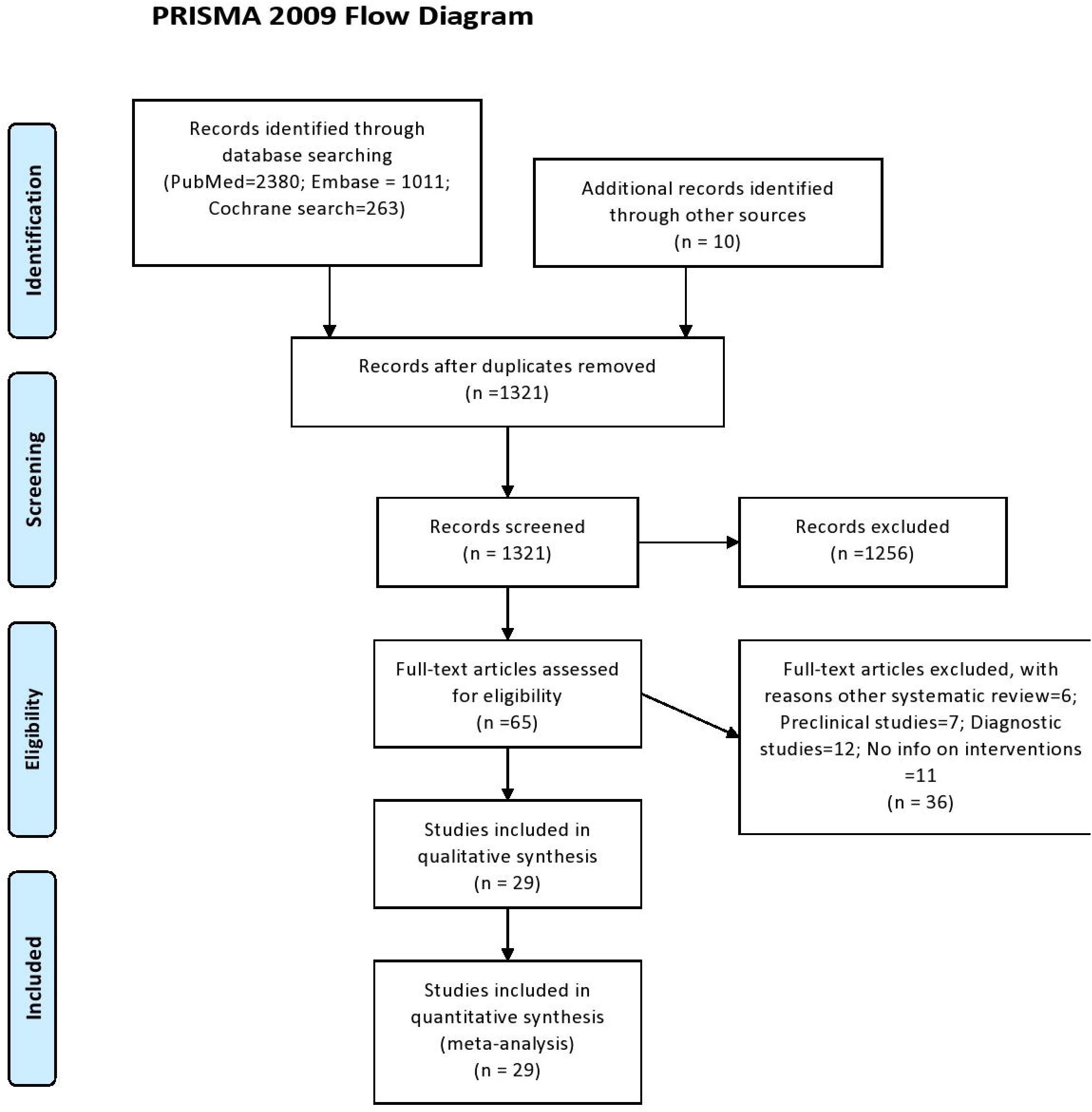
PRISMA Flow diagram

**Table 1.**
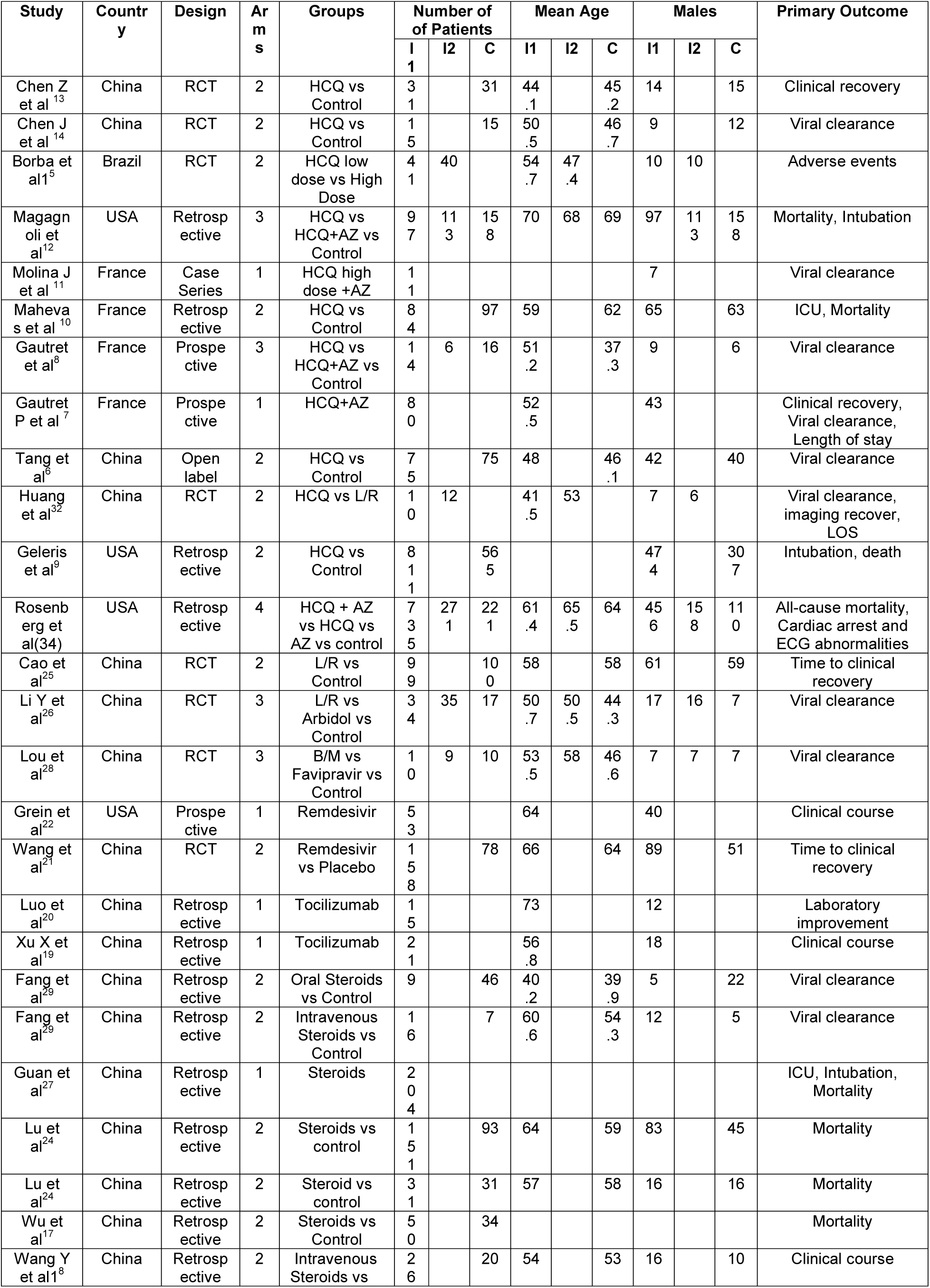
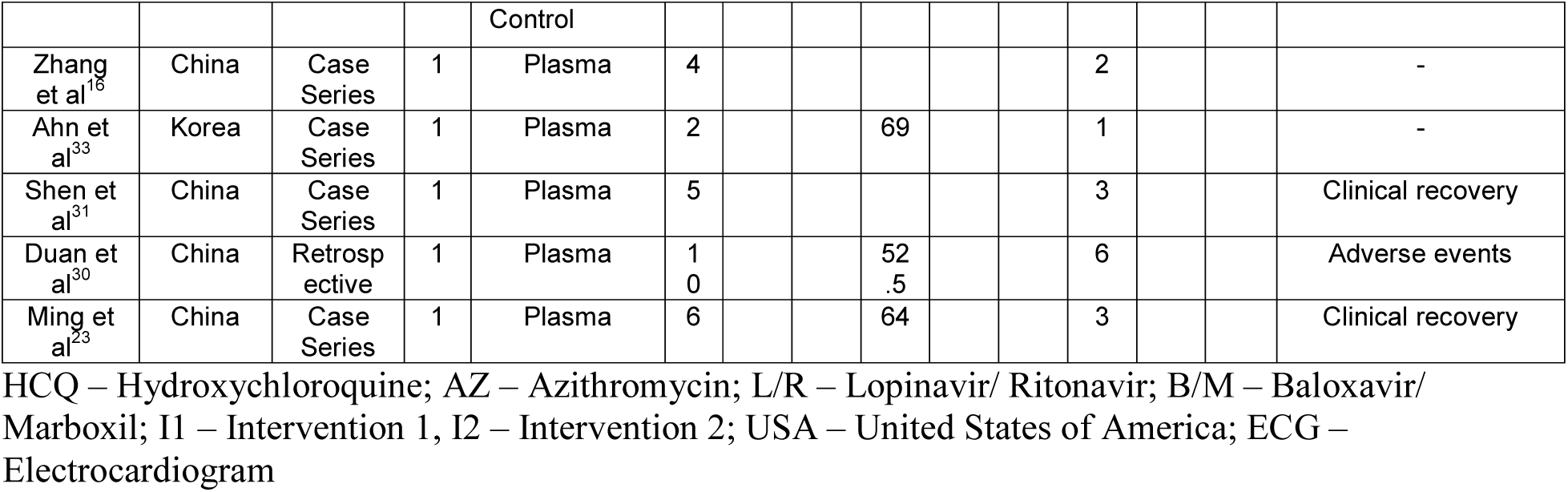
Study design, demographics, intervention arms and outcomes

### Risk of bias assessment

Eight RCTs were part of this meta-analysis. Of these 3 were at low risk of bias and 5 were at high risk. Risk of bias summary has been provided in **Appendix 4**.

### Primary outcome: All-cause mortality

Twenty-four studies provided data on mortality in the intervention arm and the pooled all-cause in-hospital mortality rate was 12.8% (95%CI: 8.1%-17.4%) for a median follow up duration of 14 (IQR: 10-18.5) days (**Table 2**). Comparing the mortality between the intervention arm and control arms, 10 studies (n = 3894) provided the data, with a pooled rate of 17.1% (95% CI: 9.1%-27.4%) in the intervention arm and 14.8% (95% CI: 9.4% - 20.1%) in the control arm, with no significant difference between the two groups (OR: 1.36, 95% CI: 0.97-1.89, I^2^ = 46%, p = 0.07) (**Figure 2A**). The NNH was calculated to be 43. When analysis was restricted to only 4 HCQ based studies (n = 3152), the mortality was significantly higher in the HCQ group (OR: 1.86, 95% CI: 1.38 – 2.50, I^2^ = 29%, p < 0.001) (NNH – 13).(**Figure 2B**) A further sub-group analysis for only 2 studies (n = 212) which used only HCQ for treatment without any other confounders like azithromycin and the mortality was still significantly higher in the HCQ group (OR: 2.17, 95% CI: 1.26-3.72, I^2^ = 43%) (NNH – 9). Comparing intervention and control arms, sub-group analysis performed for antiviral studies only (n = 550) (OR – 0.83, 95% CI: 0.49 – 1.38), steroid based studies (n = 192) (OR: 0.96, 95% CI: 0.40 – 2.31), moderate to severe disease patients (n = 2184) (OR: 1.09, 95% CI: 0.56 – 1.57), severe disease patients (n = 627) (OR: 0.87, 95% CI: 0.58-1.31) **(Appendix figure 1)** and RCTs only (n = 550) (OR: 0.83, 95%CI: 0.49 – 1.38) **(Appendix figure 2)**, did not show a statistically significant difference between the two groups **(Appendix table 1)**.

**Figure 2A.**
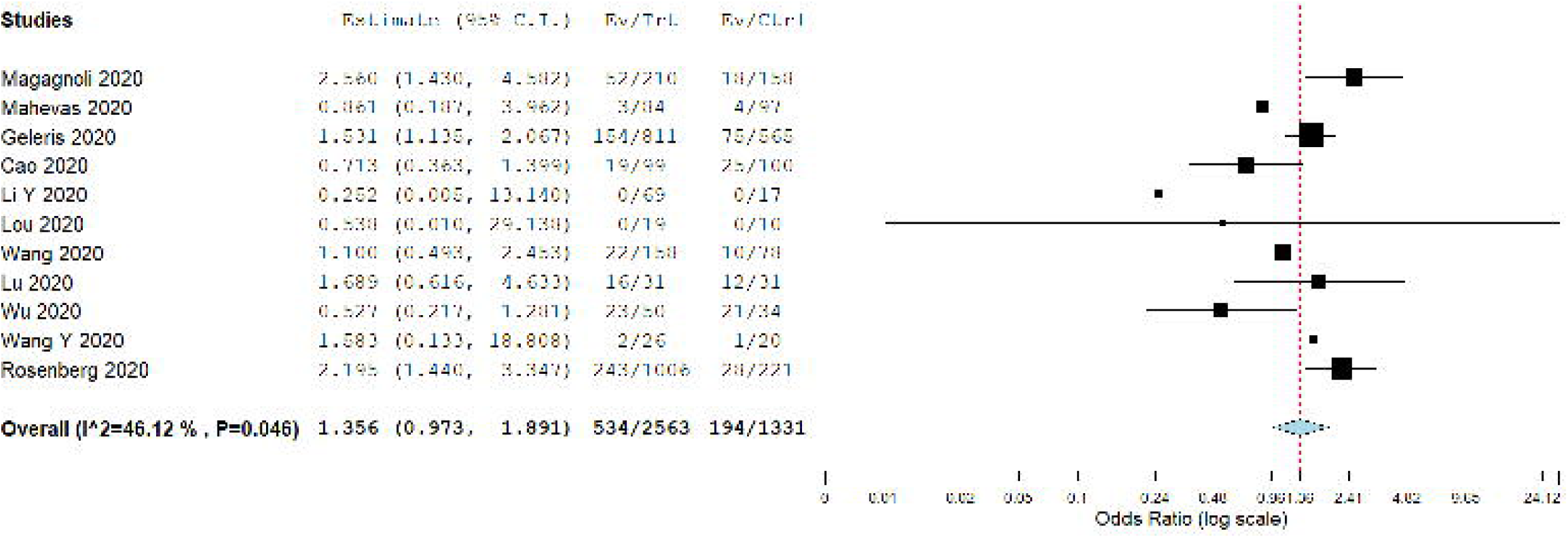
Odds ratio comparing all-cause in hospital mortality in intervention and control arms

**Figure 2B.**
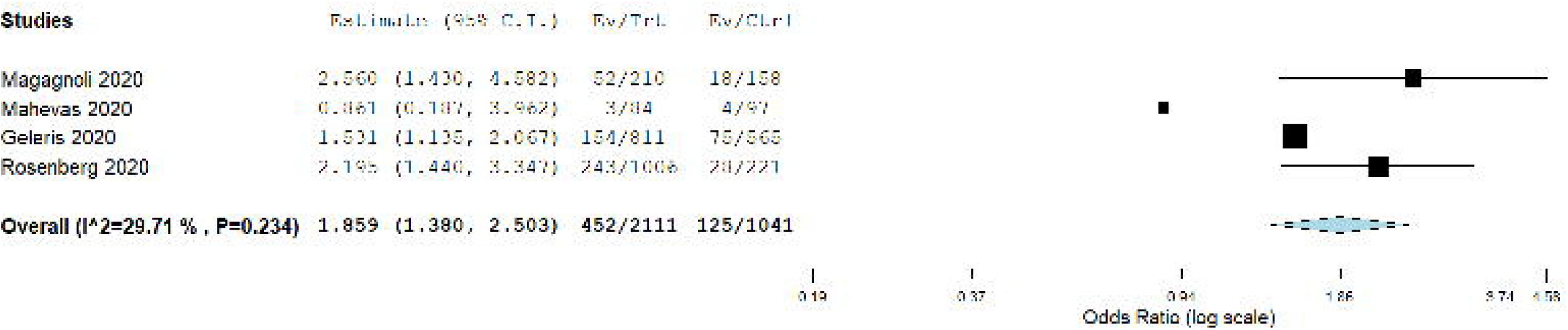
Odds ratio comparing all-cause in hospital mortality in intervention and control arms in hydroxychloroquine based studies

**Table 2.** Study outcomes in the intervention and control groups with odds ratios

| Outcome | Intervention group only | Intervention group studies common with control group | Control group only | Odds ratio |
| --- | --- | --- | --- | --- |
| All-cause mortality | 11.9 % (95% CI: 7.7%-16.1%) (n = 2071) | 16.2% (95% CI: 8.8% - 23.6%) (n = 1557) | 15.4% (95% CI: 9.1% - 21.6%) (n = 1110) | 1.22 (95% CI: 0.85 – 1.75, I<sup>2</sup> = 39%) (n = 2667) |
| Mechanical ventilation rate | 18.6% (95% CI: 10.9% - 26.3%) (n = 1456) | 13.5% (95% CI: 7% - 19.9%) (n = 1382) | 9.8% (95% CI: 4.5% - 15.2%) (n = 935) | 1.58 (95% CI: 0.60 – 4.15, I<sup>2</sup> = 85%) (n = 2317) |
| Antiviral clearance rate | 80% (95% CI: 70.7% - 89.4%) (n = 393) | 74.9% (95% CI: 59.5% - 90.3%) (n = 257) | 66.8% (95% CI: 42.6% - 91.1%) (n = 204) | 1.86 (95% CI: 0.76 – 4.54, I<sup>2</sup> = 58%) (n = 461) |
| Clinical recovery rate | 79.7% (95% CI: 78.9% - 88.4%) (n = 558) | 64.1% (95% CI: 51.5% - 76.8%) (n = 340) | 56.9% (95% CI: 43.8% - 69.9%) (n = 243) | 1.41 (95% CI: 0.99 – 2.02, I<sup>2</sup> = 0%) (n = 583) |
| Progression to severe disease | 11.6% (95% CI: 5.4% - 17.8%) (n = 387) | 13.4% (95% CI: 2.2% - 24.6%) (n = 218) | 12.8 (95% CI: 4.8% - 20.8%) (n = 168) | 1.19 (95% CI: 0.67 – 2.13, I<sup>2</sup> = 0%) (n = 386) |
| Radiological improvement rate | 86.3% (95% CI: 77.2% - 95.5%) (n = 140) | 83.6% (95% CI: 67.6% - 99.7%) (n = 107) | 82.6% (95% CI: 60.3% - 100%) (n = 60) | 1.02 (95% CI: 0.13 – 8.07, I<sup>2</sup> = 63%) (n = 167) |
| Discharge rate | 69.8% (95% CI: 60.3% - 79.3%) (n = 1374) | 68.6% (95% CI: 62.2% - 75%) (n = 1171) | 78.6% (95% CI: 67.8% - 89.4%) (n = 800) | 0.55 (95% CI: 0.29 – 1.03, I<sup>2</sup> = 81%) |
| Adverse events rate | 23.3% (95% CI: 12.1% - 34.5%) (n = 791) | 34% (95% CI: 13.9% - 54.1%) (n = 436) | 29.5% (95% CI: 9.4% - 49.6%) (n = 310) | 1.44 (95% CI: 0.70 – 2.94, I<sup>2</sup> = 62%) (n = 754) |

## Secondary outcomes

## Rate of mechanical ventilation

Nine studies (n = 1456) reported need for mechanical ventilation in patients in the intervention arm, with a pooled intubation rate of 18.6% (95% CI: 10.9% - 26.3%) (**Table 2**). Comparing the 7 studies (n = 2317) which also provided information on control population, the pooled rates in the intervention and control arms were 13.5% vs 9.8%, respectively with no significant difference between the two groups (OR: 1.58, 95% CI: 0.60 – 4.15, I^2^ = 85%) (NNT – 27) (**Figure 3A**). There was no significant difference in the outcome when analysis was restricted to HCQ and anti-viral based studies.

**Figure 3A.**
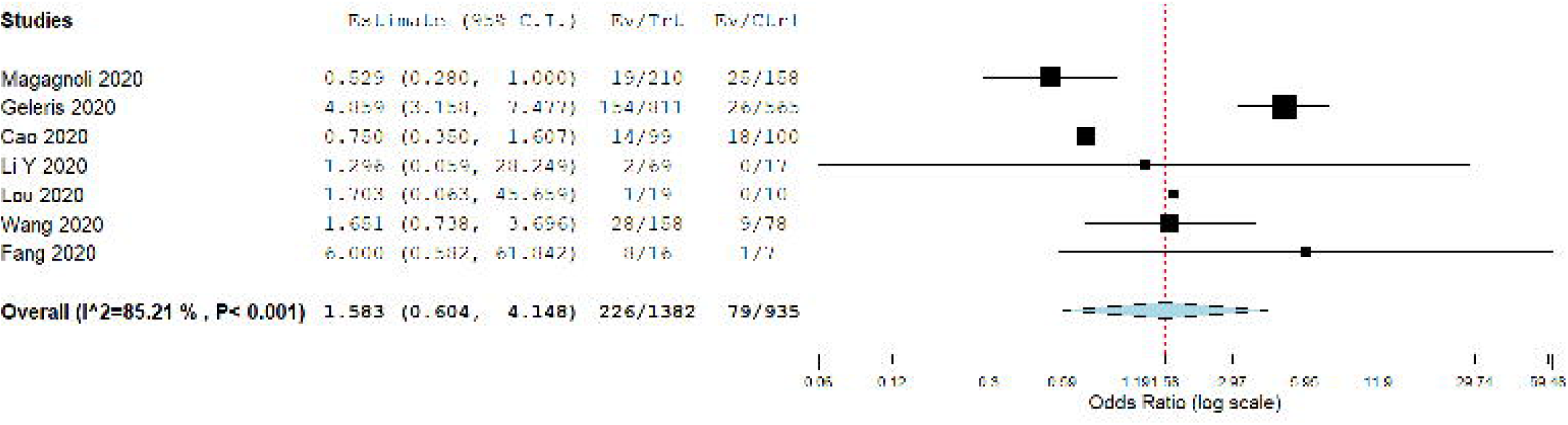
Odds ratio comparing rates of mechanical ventilation in intervention and control arms

## Viral clearance

Fifteen studies reported data on either the proportion of patients with antiviral clearance at the end of the study or the median duration for antiviral clearance. The pooled proportion of patients with antiviral clearance in the intervention arm (n = 393) was 80% (95% CI: 70.7% - 89.4%). Comparing the 6 studies (n = 461) reporting data on antiviral clearance in intervention and control groups, the pooled rates were 74.9% vs 66.8%, respectively with no significant difference between the two groups (OR: 1.86, 95% CI: 0.76 – 4.54, I^2^ = 58%) (NNT – 10) **(Appendix figure 3)**. When the analysis was restricted to HCQ based and anti-viral based studies, there was still no significant difference between the two groups. The median duration for antiviral clearance in the intervention arm (n = 308) was 6.1 (IQR: 4.3 – 8.8) days and in the control arm (n = 170) was 9 (IQR: 4.5 – 14) days, with no significant difference between the two groups (p = 0.37)

## Clinical recovery

Fourteen studies reported data on either the proportion of patients who had clinical recovery or median time to clinical recovery. The pooled rate of proportion of patients with clinical recovery in the intervention arm (n = 558) was 79.7% (95% CI: 78.9% - 88.4%). Comparing the 4 studies reporting data in intervention and control arms, the pooled rates were 64.1% and 52.8% (NNT – 9), respectively with no significant difference between the two groups (OR: 1.41, 95% CI: 0.99 – 2.02, I^2^ = 0%) (**Figure 3B**). Decrease in oxygen requirements in both groups was reported in 2 studies (n = 375), with no significant difference between both the groups (OR: 1.05, 95% CI: 0.65 – 1.71, I^2^ = 3%).

**Figure 3B.**
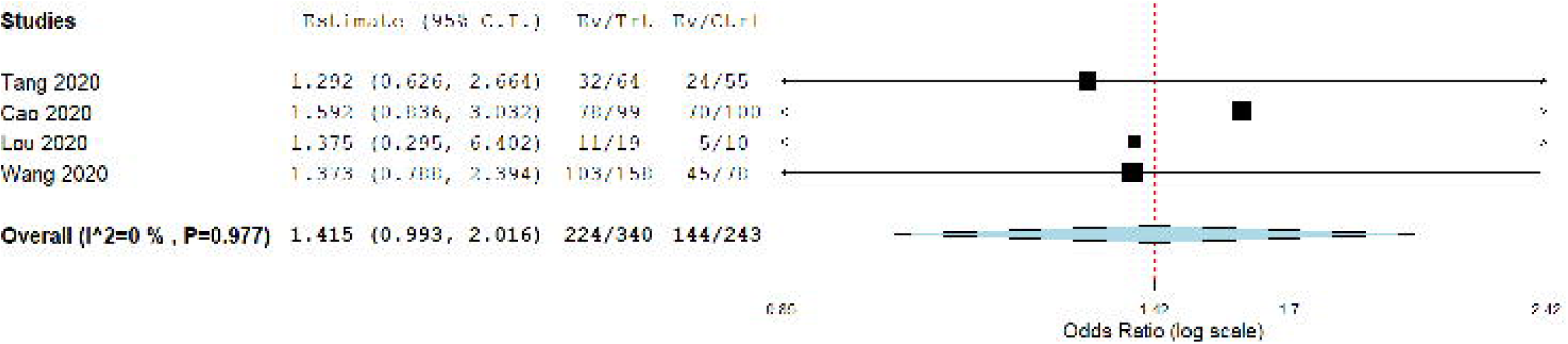
Odds ratio comparing clinical recovery rates in intervention and control arms

The median time to clinical recovery was 14 (IQR: 8.2 – 19) days in the intervention group (n = 451) and 16 (IQR: 14.3 - 22) days in the control group (n = 263), with no significant difference between the two groups (p = 0.25)

## Progression to severe disease

Nine studies reported data on worsening of clinical status in the hospital in mild-moderate severity patients, with a pooled rate of 11.6% (95% CI: 5.4% - 17.8%) in the intervention arm (n = 387) over a median duration of 13 (IQR: 9.5 – 19.5) days. Comparing the pooled rates in five studies reporting the outcome in both groups (n = 386), the pooled rates were 13.4% and 12.8% in the intervention and control groups, respectively with no significant difference between the two groups (OR: 1.19, 95% CI: 0.67 – 2.13, I^2^ = 0%). Sub-group analysis restricted to HCQ based and antiviral studies also did not reveal any significant difference.

## Adverse events

Sixteen studies (n = 791) reported the rate of adverse events in the intervention group with a pooled adverse event rate of 23.3% (95% CI: 12.1% - 34.5%). Six studies (n = 754) compared intervention and control groups with pooled adverse event rates of 34% and 29.5% respectively, with no significant difference between the two groups (OR: 1.44, 95% CI: 0.70 – 2.94) (NNT – 22) **(Figure 4A)**. On sub-group analysis, the incidence of adverse events was significantly higher in the HCQ group (OR: 3.88, 95% CI: 1.60 – 9.45, I^2^ = 0%) **(Figure 4B)** with a NNH of 7, but there was no significant difference for studies with antiviral agents.

**Figure 4A.**
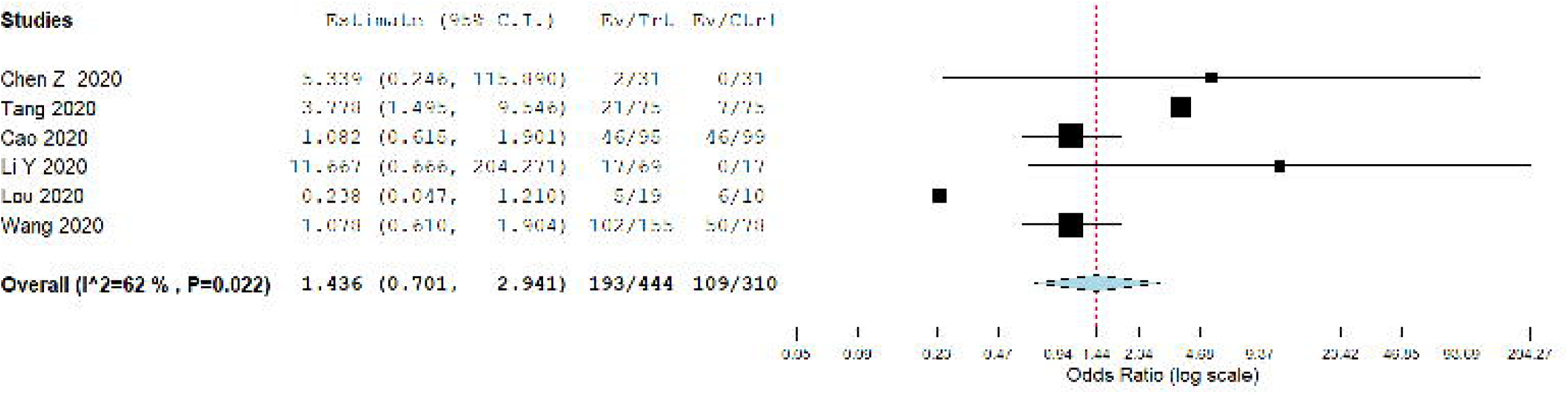
Odds ratio comparing adverse events rates in intervention and control arms

**Figure 4B.**
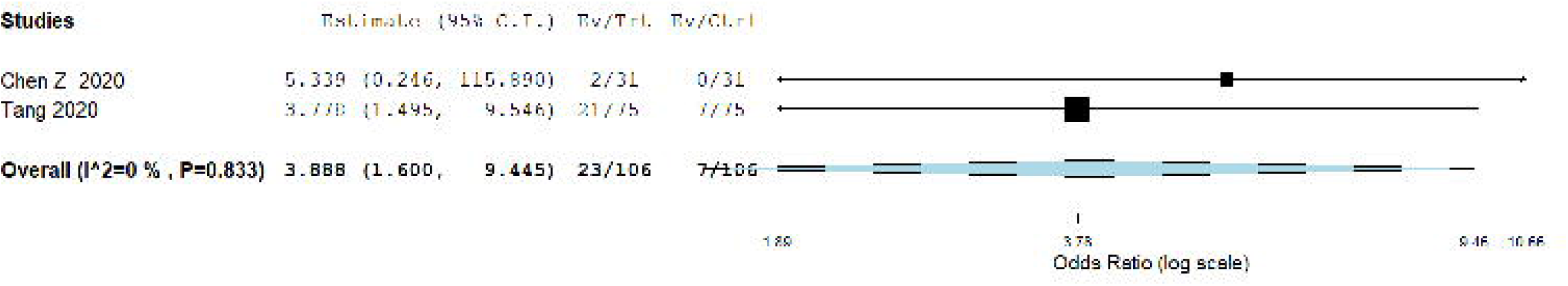
Odds ratio comparing adverse events in intervention and control arms in HCQ based studies

There was also no significant difference between both the groups in radiological improvement and discharge rates. The individual pooled rates and ORs are provided in **Table 2**. Sub-group analysis for only RCTs for available outcomes are provided in **Appendix table 2**.

## Discussion

In this systematic review and meta-analysis of 5207 patients from 29 studies, pooled outcome for any therapeutic intervention including HCQ, Remdesivir, Lopinavir/Ritonavir, Steroids, Tocilizumab, Convalescent plasma therapy did not show a survival benefit compared to control arms. HCQ use was associated with significantly increased all-cause inpatient mortality and adverse event rates. There were no significant benefits with any therapeutic intervention in changing the natural history of SARS-CoV-2 infection assessed in terms of rate of mechanical ventilation, viral clearance, time to discharge, time to clinical recovery and radiological improvement.

HCQ increases the endosomal pH and prevents fusion of the host membrane with SARS-CoV-2 thereby interfering with the viral replication cycle. Several pre-clinical studies showed invitro activity against SARS-CoV-2 leading to clinical use of HCQ in COVID-19 disease(35, 36). An initial case series of 26 patients from France comparing HCQ and control groups suggested that HCQ leads to rapid anti-viral clearance in 70% of patients compared to 12.5% in controls. This study was fraught with methodological inconsistencies like enrollment of asymptomatic individuals, omission of 6 patients from analysis (HCQ patients of whom one died and 3 were transferred to ICU)(9). In a randomized study of 62 patients from China, patients treated with HCQ showed radiological improvement in resolution of lung lesions as well as reduction in clinical progression of disease. The commonality of the initial studies on HCQ were relatively small sample size, inappropriate control groups, lack of clarity in defining the study outcomes(14). Two larger prospective RCTs from France and Brazil show that HCQ/chloroquine use is associated with increased incidence of cardiac events with no survival benefit(12, 16). In view of conflicting data outcomes, the National Institute of Health (NIH), USA recommends that there is insufficient clinical data to recommend either for or against using chloroquine or hydroxychloroquine for the treatment of COVID-19(37).Despite these issues, HCQ is currently one of the most commonly used medications in various parts of the world. Our analysis shows that HCQ based regimens had increased rate of mortality (NNH – 13) and adverse events (NNH – 7) compared to control patients. We hope our meta-analysis adds more evidence to dampen its use in view of lack of benefit and increased side effects.

Remdesivir was originally designed for use against Ebola. Remdesivir was studied in severe COVID-19 in China showed that the drug did not show any benefit in terms of time to recovery as well as 28 day mortality outcome, though the study was terminated prematurely in view of difficulty in patient accrual(22).ACTT NIH study showed that Remdesivir accelerated the time to recovery to 11 days compared to 15 days in the placebo arm with no mortality benefit. This prompted an emergency Food and Drug Ddministration (FDA) authorization for use in COVID-19 patients. There are calls for Remdesivir to be taken as the standard of care control in future clinical studies. There have been questions raised about the NIH study due to limitations such as change in the primary endpoint of study after initiation of the trial, lack of mortality benefit; study in moderate disease patients who tend to recover spontaneously by the end of second week(38).There are ethical concerns among the scientific community that drugs without proven mortality benefit or reduction in the need for ventilatory support may be promoted in view of aggressive pharmaceutical lobbying. The current pandemic rings echo bells of the 2009 H1N1 pandemic and the desperate stockpiling of oseltamivir, whose proclaimed efficacy was claimed to be a byproduct of concealed information and aggressive lobbying by pharmaceutical companies(39) The published or preprint data for other drugs like Favipravir, Baloxavir/ Marboxil, corticosteroids, convalescent plasma is currently insufficient to make any specific recommendation and our meta-analysis also suggests the same.

Our meta-analysis shows that none of the so far studied interventions have a tangible benefit to change the course of disease outcomes with the current published evidence. The clinical studies that compare various interventions like the World health organization (WHO) sponsored solidarity trial that compares Remdesivir, chloroquine or hydroxychloroquine, lopinavir plus ritonavir, and interferon-beta with control arm and has all-cause mortality as the primary outcome is the need of the hour and the results are eagerly awaited(40).

The strengths of our study are as follows: We included 29 studies with more than 5200 patients in our analysis with various interventions. Our review is extensive, by including the available interventions and providing clinically relevant outcomes in comparison with controls. Several sub-group analyses were also performed based on study interventions and design. Heterogeneity in most of our outcomes was mild to moderate but we performed sub-group analysis in RCTs to further reduce the heterogeneity.

### Limitations

Our study has several limitations. The study design, patient population and the outcomes assessed were variable in different studies. Even though the intervention arms were clearly defined in most studies, some of the patients in those arms also received other medications and outcomes for such patients could not be excluded separately, which could have confounded the results. Different levels of disease severity of patients on study entry could lead to heterogeneity in outcomes but we tried to address it by performing sub-group analyses based on disease severity for outcomes when possible. Duration of follow-up was variable across studies and entire patient data at the end of study may not have been represented which is a limitation of the published literature. Adverse events reported include medication related adverse events and also symptoms in both groups, which could be related to SARS-CoV-2, but this was unanimously reported across all studies. The dose of medications, especially HCQ, was variable in studies and dose based analysis could not be performed. Data from pre-published studies were also included in our analysis but we had included them to provide a more comprehensive overview to prevent misinterpretation of results to the best of our capabilities. The results of our study should hence be interpreted with caution keeping these limitations in mind.

## Conclusions

In this meta-analysis, there was no overall mortality or clinical benefit for most therapeutic interventions but the use of HCQ was associated with increased mortality rates and increased risk of adverse events in SARS-CoV-2 patients. None of the other therapeutic interventions like Lopinavir/Ritonavir, Remdesivir, Tocilizumab seemed to alter the natural clinical course of the disease based on the available literature. There is a need for well-designed randomized clinical trials to further investigate the efficacy and safety of various therapeutic interventions.

## Data Availability

All data in the manuscript is original and was created by the authors involved in this manuscript

## Abbreviations

SARS-CoV-2 -: Severe Acute Respiratory Syndrome related Coronavirus-2
HCQ –: Hydroxychloroquine
COVID-19 –: Coronavirus disease 2019
PRISMA –: Preferred Reporting Items for Systematic Reviews and Meta-Analyses
RCT –: Randomized Controlled Trials
NNT –: Number needed to treat
NNH –: Number needed to harm
IQR –: Interquartile range
CI –: Confidence interval
OR –: Odds ratio
NIH –: National Institute of Health
FDA –: Food and Drug Administration
WHO –: World Health Organization

## Author approval

All authors have seen and approved the submission of this manuscript

## Competing interests

None

## Declarations

None

## Funding

None

## Acknowledgements

None

## Disclaimer

None

## Author bio

Dr. Thoguluva Chandrasekar is currently a Gastroenterology fellow at the University of Kansas Medical Center. His research interest is in the field of Gastroenterology, primarily on Barrett’s esophagus and colorectal cancer. He will be specializing in Advanced Endoscopy training soon. Given the global pandemic due to SARS-CoV-2 infection, his interest has also been to investigate the GI manifestations in SARS-COV-2 infection and effect of various therapeutic interventions in SARS-CoV-2.

## Appendix legends

**Appendix 1** –PRISMA checklist

**Appendix 2** –Medical Subject Entrée Terms used for search

**Appendix 3** –Definition of outcomes

**Appendix 4** –Risk of Bias summary for RCTs

**Appendix table 1** –All cause mortality for various sub-groups

**Appendix table 2** –Outcomes only for Randomized Controlled Trials

**Appendix figure 1 –Odds ratio comparing all-cause in hospital mortality in the interventions and control arms with severe disease**

**Appendix figure 2 - Odds ratio comparing all-cause in hospital mortality in the intervention and control arms for randomized controlled trials only**

**Appendix figure 3 –Odds ratio comparing viral clearance rates in intervention and control arms**

---

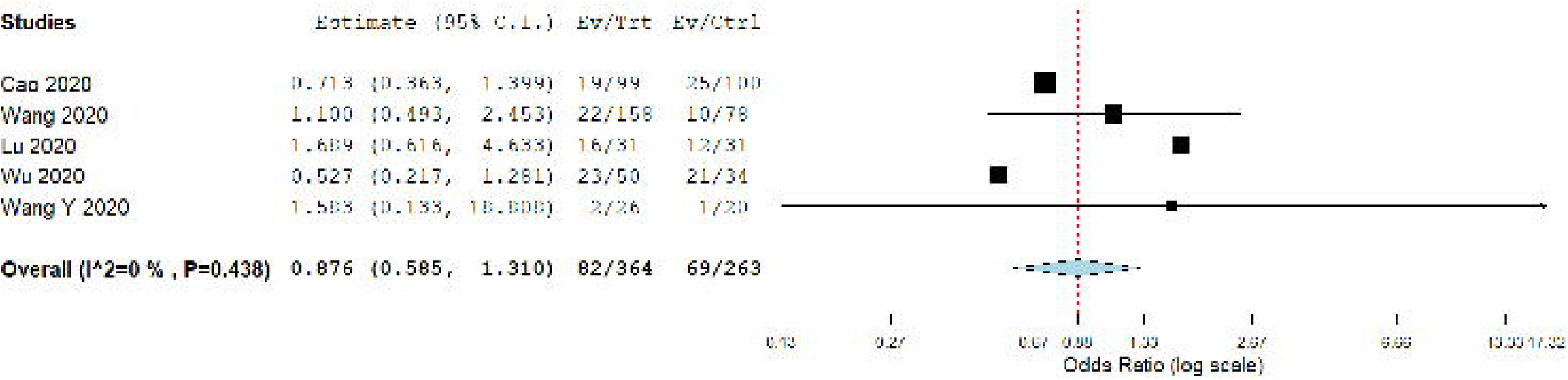

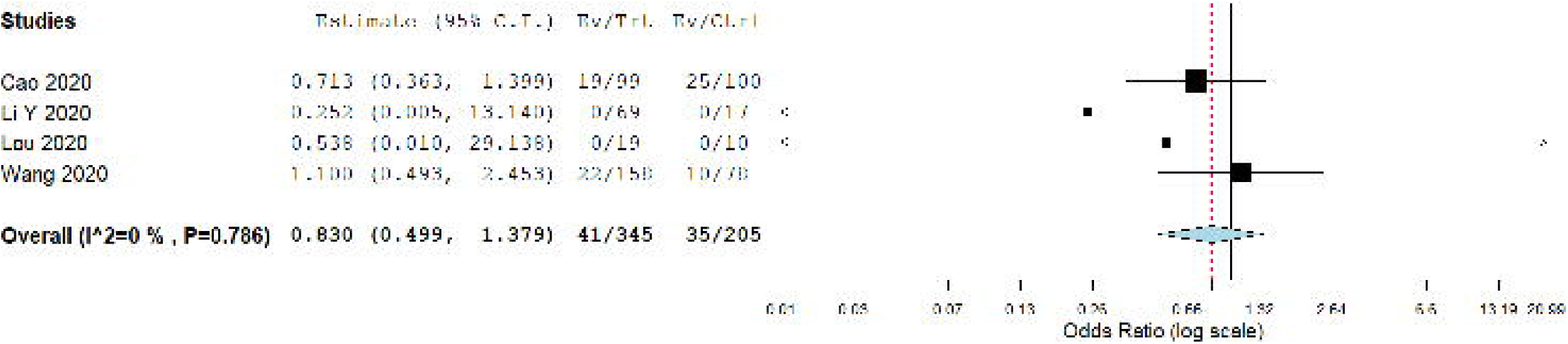

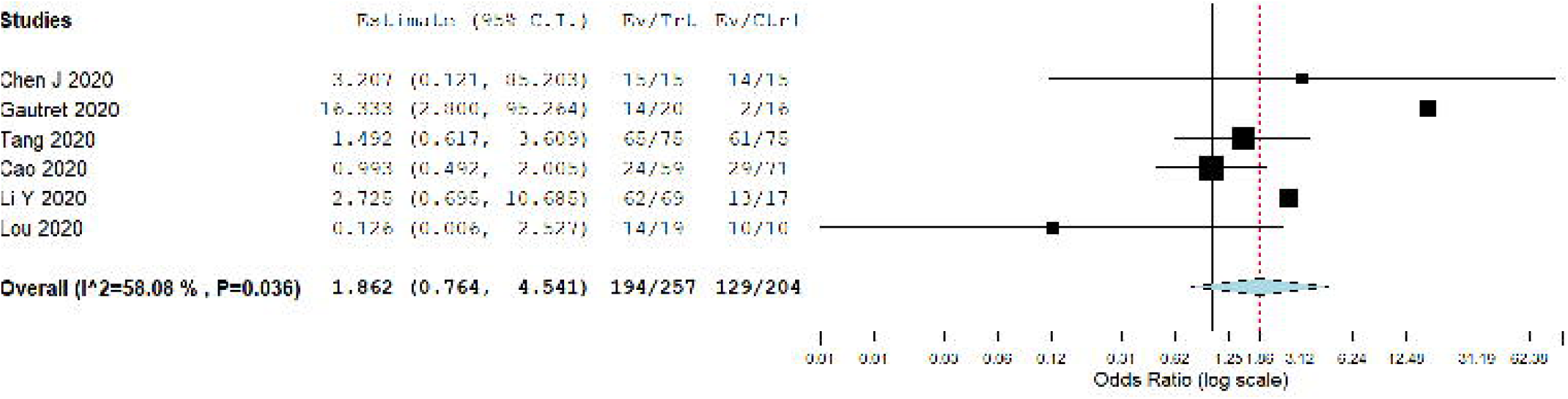

## Notes

### Competing Interest Statement

The authors have declared no competing interest.

### Funding Statement

No source of funding to declare

### Author Declarations

No IRB approval was needed for this study as this is a systematic review and no patient information was disclosed

